# Parkinson’s Disease Reshapes Gut Microbiota Stochasticity and Composition: A Near-Neutral Modeling and Network Analysis

**DOI:** 10.1101/2025.03.16.25324047

**Authors:** Yuting Qiao, Zhanshan (Sam) Ma

**Affiliations:** Computational Biology and Medical Ecology Lab Kunming Institute of Zoology, Chinese Academy of Sciences, Kunming, China; Microbiome Medicine and Advanced AI Lab, Kunming, China; Faculty of Arts and Science Harvard University Cambridge, MA, 20138, USA

**Keywords:** Parkinson’s Disease (PD), Sloan’s near-neutral model (SNM), Neutral theory, Network analysis, Gut microbiome

## Abstract

Parkinson’s disease (PD) is increasingly linked to gut microbiome dysbiosis, yet the ecological mechanisms driving these microbial changes remain poorly understood. This study integrates Sloan’s near-neutral model (SNM), the normalized stochasticity ratio (NSR) framework, and ecological network analysis to investigate how PD reshapes gut microbiota stochasticity, composition, and interactions. Using eight publicly available gut microbiome datasets (1,957 samples: 804 healthy, 1,153 PD), we applied SNM to classify species into neutral, positively selected, and negatively selected categories. While the proportions of these categories remained unchanged between healthy and PD groups (neutral: ∼40%, positively selected: ∼47%, negatively selected: ∼13%), NSR analysis revealed holistic alterations in stochasticity due to PD, suggesting increased stochastic drifts. Shared species analysis (SSA) reconciled the apparent inconsistency between SNM and NSR by demonstrating significant compositional shifts within each species category (P < 0.05). Network analysis showed that neutral species had fewer antagonistic interactions (higher positive-to-negative link ratios) compared to selected species, consistent with their ecological equivalence, while negatively selected species dominated in relative abundance across both groups, underscoring their ecological significance. Taylor’s Power Law Extensions (TPLE) indicated invariant heterogeneity-scaling across species categories and disease status, reflecting consistent spatial aggregation patterns. Collectively, these findings reveal that PD reshapes gut microbiota through altered stochasticity and species interactions rather than changes in the overall structure of species categories. This study provides novel ecological insights into PD-associated microbial dysbiosis, highlighting the interplay of stochastic and deterministic processes in microbial community assembly, with potential implications for identifying microbial biomarkers and therapeutic targets.

## Introduction

The human gut microbiota, a complex ecosystem comprising trillions of microorganisms, plays a pivotal role in modulating host physiology, including immune regulation, nutrient metabolism, and neural signaling (Lynch & Pedersen, 2016; Cryan et al., 2020). Emerging evidence highlights its dysbiosis as a potential contributor to neurodegenerative disorders, particularly Parkinson’s disease (PD) (Sampson et al., 2016; Mayer et al., 2014). While PD is classically characterized by motor symptoms linked to dopaminergic neuron loss, recent studies reveal that gastrointestinal dysfunction and microbial alterations often precede neurological manifestations, suggesting a gut-brain axis involvement (Braak et al., 2003; Forsyth et al., 2011). Despite growing recognition of microbial compositional shifts in PD patients (Keshavarzian et al., 2015; Qian et al., 2018), the ecological mechanisms driving these changes—whether deterministic selection (e.g., host physiology, diet) or stochastic processes (e.g., random dispersal, drift)—remain poorly resolved. This knowledge gap limits our understanding of the etiologic role of microbiota in the pathogenesis of Parkinson’s disease.

Traditional analyses of gut microbiota in PD have focused predominantly on taxonomic differences, identifying reduced abundances of anti-inflammatory taxa (e.g., *Prevotella, Faecalibacterium*) and enrichment of pro-inflammatory species (e.g., *Enterobacteriaceae*) (Houser et al., 2017; Scheperjans et al., 2015). However, such descriptive approaches fail to address the ecological forces shaping community assembly. Neutral theory, originally developed for macro-ecosystems, posits that species coexistence arises primarily from stochastic birth-death dynamics and immigration rather than niche specialization (Hubbell, 2001). Sloan et al. (2006, 2007) extended this framework into a “near-neutral” model (SNM), which relaxes strict neutrality by allowing species-specific fitness differences, thereby bridging deterministic selection and stochastic drift. Applied to microbial communities, SNM quantifies the relative contributions of neutral processes (e.g., random turnover) versus selective pressures (e.g., host immune filtering), offering mechanistic insights into community assembly (Zhou & Ning, 2017).

Microbial communities in human gut environments are structured by complex interactions between stochastic (neutral) and deterministic (niche-related) ecological processes (Hubbell, 2001; Vellend, 2010). Recent ecological modeling frameworks, particularly those based on neutral theory, offer powerful methods to quantify and differentiate the roles of deterministic selection (niche processes) versus stochastic processes in shaping community composition (Hubbell, 2001). Hubbell’s neutral theory (HNM) has provided foundational insights by assuming functional equivalence among species; however, strict neutrality is rarely supported empirically, particularly in host-associated microbiomes (Zhou et al., 2017). To overcome these limitations, Sloan et al. developed a near-neutral model (SNM), an extension of Hubbell’s model that relaxes strict neutrality assumptions, allowing for subtle competitive advantages or disadvantages among species (Sloan et al., 2006, 2007). This model offers a valuable ecological framework for interpreting microbial community assembly at a finer resolution, distinguishing between neutral, positively selected, and negatively selected species through comparison of observed abundances against model predictions (Sloan et al., 2006, 2007). Yet, its application to disease-associated dysbiosis, particularly in neurodegenerative contexts, remains unexplored.

Moreover, traditional methods analyzing microbial communities have primarily emphasized deterministic explanations, largely overlooking stochastic processes. Recently, Ning et al. introduced the normalized stochasticity ratio (NSR) framework as a robust tool for quantifying the extent of stochastic versus deterministic influences within microbial community assemblies (Ning et al., 2019). The NSR framework evaluates whether community structure deviates significantly from neutral expectations, effectively delineating the upper limits of community stochasticity. Such insights are critical, as elevated stochasticity might indicate disrupted ecological selection processes, suggesting microbial instability or susceptibility to perturbations linked to diseases such as PD (Ning et al., 2019).

Meanwhile, community assembly processes also significantly influence the co-occurrence and interaction networks within microbiomes. Ecological network analysis is increasingly employed to identify potential microbial interactions, keystone taxa, and ecosystem stability indicators in health and disease states (Faust & Raes 2012). Network analysis has emerged as a powerful tool to unravel microbial interaction patterns, revealing keystone species and functional modules that may underpin ecosystem stability or collapse (Faust et al., 2012; Berry et al., 2014). Through these co-occurrence networks, subtle yet significant changes in species interactions and connectivity may elucidate ecological disruptions associated with disease onset and progression (Banerjee et al., 2018). Integrating neutral model-based approaches with network analyses thus offers an integrative perspective on both individual species-level responses and broader microbial interaction patterns altered by diseases, providing holistic insights into microbial ecological dynamics. In PD, disrupted microbial co-occurrence networks have been reported, but whether these disruptions reflect altered deterministic interactions or amplified stochasticity is unknown (Wallen et al., 2021; Hill-Burns et al., 2017). Integrating near-neutral modeling with network analysis could disentangle these dynamics, clarifying how PD reshapes both microbial composition and ecological architecture.

In this study, we integrate Sloan’s near-neutral model (SNM) with the normalized stochasticity ratio (NSR) framework and network analyses to comprehensively investigate how PD reshapes the gut microbiota at both OTU and community levels. Specifically, we aim to (i) determine the relative importance of stochasticity and deterministic factors in shaping gut microbial communities in PD, (ii) identify microbial taxa that significantly deviate from neutral expectations, thus highlighting potential PD-associated selective processes, and (iii) explore alterations in microbial co-occurrence networks to uncover ecological interactions associated with PD pathogenesis. By combining these ecological frameworks and network analyses, we anticipate revealing novel ecological insights into PD-associated microbial dysbiosis, potentially identifying microbial markers for disease monitoring, diagnosis, or intervention. This integrative approach not only advances our understanding of the gut-brain axis in PD but also provides a methodological framework for studying microbial ecology in other neurodegenerative diseases.

## Material and Methods

### Bioinformatics Analysis of Gut Microbiomes Associated with Parkinson’s Disease (PD)

This study reanalyzed eight publicly available human gut microbiome datasets associated with Parkinson’s disease, comprising a total of 1,957 samples—804 from healthy individuals and 1,153 from PD patients (see Table S1 for details). Raw sequencing data were obtained from the NCBI database (https://www.ncbi.nlm.nih.gov/) and processed using a bioinformatics pipeline that included Trimmomatic (v0.39) for adapter trimming and quality filtering, Kraken2 (v2.1.2) for taxonomic classification, and Bracken (v2.6) for abundance estimation, utilizing the 16S Greengenes database (March 25, 2020). KrakenTools and R software (version 4.1.2) were employed to convert the data into a usable format. A summary of the datasets and operational taxonomic unit (OTU) tables is provided in Table S1.

### Sloan’s Near-Neutral Model (SNM) and Fisher’s Exact Test for Neutrality Analysis

Sloan et al. (2006, 2007) proposed a near-neutral model (SNM) based on Hubbell’s neutral theory, which considers two types of communities: a source community and a local community, analogous to the “mainland” and “island” in island biogeography theory. Unlike Hubbell’s strict neutral model, SNM relaxes the assumption of neutrality by allowing species to exhibit competitive advantages (positive selection) or disadvantages (negative selection). This flexibility makes SNM particularly well-suited for analyzing large prokaryotic communities without relying on observed species abundance distributions.

In the model, the local community is assumed to be saturated with individuals. When an individual dies or leaves, its vacancy is filled through one of two processes: with probability *m*, an immigrant from the source community replaces the lost individual, or with probability (1 − *m*), the vacancy is filled by the offspring of a randomly selected local individual. The probabilities for the abundance of the *i*-th operational taxonomic unit (OTU) to increase by one, decrease by one, or remain unchanged are defined as:

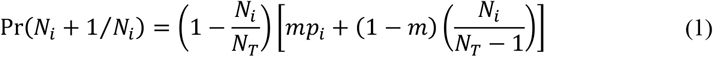

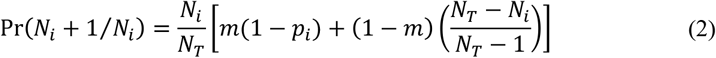

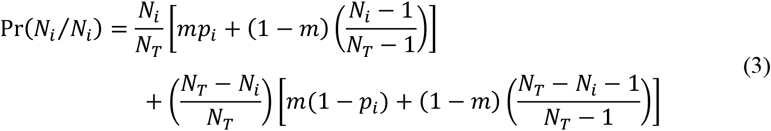

Here, *p*_*i*_ denotes the occurrence frequency of the *i*-th OTU in the source community, *N*_*i*_ represents its abundance in the local community, and *x*_*i*_ = *N*_*i*_/*N*_T_ defines its local occurrence frequency. The model uses these probabilities to assess the neutrality status of each species. Specifically, if the observed *x*_*i*_ falls within the 95% confidence interval predicted by the model, the species is considered neutral. If *x*_*i*_ exceeds the upper bound, it is classified as above neutral (positively selected), indicating a competitive advantage, while if *x*_*i*_ falls below the lower bound, it is deemed below neutral (negatively selected), reflecting a competitive disadvantage. Sloan’s model is particularly valuable for species-level assessments and addresses sampling limitations inherent in metagenomic studies. Although it does not provide a community-level test statistic like Hubbell’s model, its goodness-of-fit can be evaluated using a generalized *R*^*2*^ metric. For further details on the model, readers are referred to Sloan et al. and Burns et al.

Here, we applied Sloan’s Near-Neutral Model (SNM) to analyze the niche-neutrality continuum of OTUs in human gut microbiome datasets associated with Parkinson’s disease (PD). The model was fitted by treating each sample site as both the source and destination community. To compare species categories between healthy (H) and diseased (D) groups, we employed Fisher’s exact test and calculated the odds ratio (OR) to quantify the strength of associations. The analysis was based on a 2×2 contingency table, structured as follows:

|  | Total number of<br>species | Number of<br>neutral/selected species |
| --- | --- | --- |
| Health | $a$ | $b$ |
| Disease | $c$ | $d$ |

Here, *a* represents the total number of species in the healthy group, and *b* denotes the total number of species in the diseased group. *c* corresponds to the number of neutral (or selected) species in the healthy group, while *d* represents the number of neutral (or selected) species in the diseased group. *N* is the total number of species across both groups. Under the null hypothesis of no association between species counts and health status, the probability of observing this specific contingency table is determined by the hypergeometric distribution:

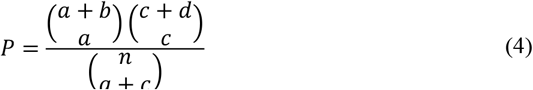

To quantify the strength and direction of the association, we calculated the odds ratio (OR), which is defined as:

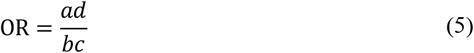

The odds ratio (OR) compares the odds of species counts in the healthy group relative to the diseased group. An OR greater than 1 indicates a positive association, suggesting higher odds of species presence in the healthy group, while an OR less than 1 reflects a negative association, implying lower odds of species presence in the diseased group.

### Normalized Stochasticity Ratio (NSR) Analysis for neutral and selected species in SNM

Ning et al. (2019) introduced the normalized stochasticity ratio (NSR) framework to quantify the upper bounds of stochasticity in community assembly. The framework is based on the premise that deterministic processes cause communities to deviate from neutral expectations, becoming either more similar or more dissimilar than predicted by a null model.

A key metric in this framework is the Ružička similarity index, a species-abundance-based extension of the Jaccard binary similarity coefficient. Let *C*_*ij*_ denote the observed similarity between the *i*-th and *j*-th communities, which is calculated as:

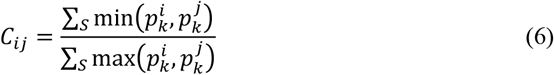

where *S* is the total number of species, and 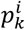 and 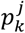 are the relative abundances of the *k*-th species in communities *i* and *j*, respectively.

Assuming there are *m* local communities within a metacommunity, let *C*_*ij*_ be the observed similarity between any two communities *i* and *j*. Denote by *E*_*ij*_ the null expected similarity for these communities in a single simulated metacommunity, and let 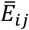 represent the average expected similarity computed over 1,000 simulated metacommunities.

There are two scenarios in evaluating community stochasticity: (*i*) When deterministic processes drive communities to be more similar: In this case, 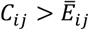,and the corresponding stochasticity ratio (type 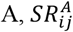) is calculated accordingly.

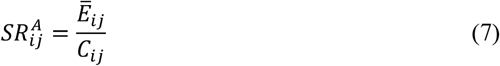

(*ii*) When deterministic processes drive communities to be more similar: In this case, 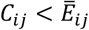,and the corresponding stochasticity ratio (type 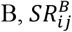) is calculated accordingly.

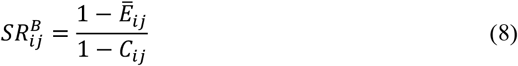

The overall stochasticity ratio for the metacommunity is then obtained by combining these pairwise ratios:

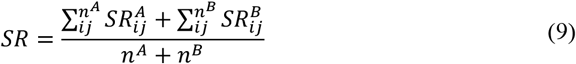

where *n*^*A*^ is the number of pairwise comparisons for which 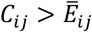 (i.e., communities are more similar than expected), and *n*^*B*^ is the number of comparisons for which 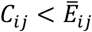 (i.e., communities are less similar than expected). The *SR* value, which ranges from 0% to 100%, represents the strength of stochasticity in community assembly; a value of 0% indicates entirely deterministic assembly, while 100% suggests complete stochasticity.

Ning et al. noted that when the expected stochasticity is very low, the *SR* may overestimate the true level of stochasticity. To address this, they proposed normalizing the stochasticity ratio. The resulting normalized stochasticity ratio (NSR) has been shown to provide a more precise estimate than the unnormalized *SR*. In this study, we adopt the NSR framework as outlined in Ning et al. to evaluate the stochasticity of species categories in SNM. The Wilcoxon test was then used to compare the differences in NSR values between healthy and diseased treatments for species categories in the SNM.

### Shared Species Analysis (SSA) for neutral and selected species in SNM

To evaluate the expected number of shared species under the null hypothesis (H_0_) across species categories in healthy and diseased groups, we applied two randomization procedures developed by Ma et al. (2019). The first, the “read randomization” algorithm, pools all sequencing reads from each treatment and randomly reassigns each read to one of the treatment cohorts, generating the expected number of shared species. The second, the “sample randomization” algorithm, treats each microbiome sample as a unit, randomly redistributing samples into pseudo-groups and aggregating reads within each pseudo-group to estimate shared species.

The expected number of shared species was taken as the average of 1000 stimulations in the randomization test (Ma 2019). Let D be the number of times the number of shared species in 1000 random samplings (i.e., random redistributions) exceeds the observed number of shared species. The pseudo p-value is calculated as follows:

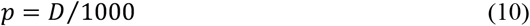

A *p*-value less than 0.05 indicates that the observed difference in shared species between healthy and diseased individuals is unlikely to be due solely to random variation, thereby suggesting a statistically significant difference between the groups.

### The species dominance network (SDN) analysis for neutral and selected species in SNM

We used species dominance *D*_*s*_ to perform dominance network analyses. Firstly, we defined species dominance (*D*_*s*_) as:

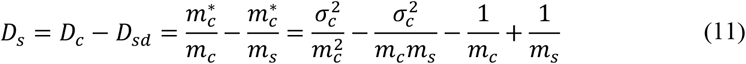

where *D*_*c*_ is community dominance, 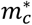 is community mean crowding, *m*_*c*_ is the mean of the population abundances (size) across all species (i.e., per species) in the community, and 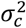 is corresponding variance and *m*_*s*_ is the population abundance (size) of the focal species of interest (*s*) in the community.

*D*_*s*_ is species specific and measures the dominance of a particular (focal) species in a community; the more dominant species there are, the larger the *D*_*s*_ value.

We constructed SDNs using the Spearman rank correlation coefficient, which was calculated between species-specific species dominance indices, *D*_*s*_ (Eq. 11). We performed standard correlation network analyses using the rcorr function of the Hmisc package in the R statistical software environment (R version 4.4.1) and Cytoscape (v3.10.3) software (Shannon et al., 2003) (Hmisc is available online). The former was used to calculate the Spearman rank correlation coefficient (*r*) between species.

### Taylor’s power law extensions (TPLE) to measure the community spatial heterogeneity for each species category in SNM

Taylor’s Power Law (TPL), introduced by L. R. Taylor (1961), describes the relationship between mean population abundance (*m*) and its variance (*V*) using a power function:

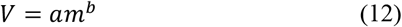

where *a* and *b* are TPL parameters. Parameter *b* is considered the heterogeneity scale parameter because it represents the slope of the variance-mean (*V*-*M*) relationship plotted on a logarithmic scale, i.e.

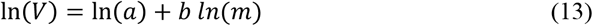

When *b*>1, the population spatial distribution is clustered (heterogeneous), when *b*=1, it is random, and when *b*<1, it is uniform or regular. The concept of population aggregation critical density (PACD) or *m*_0_, expressed as

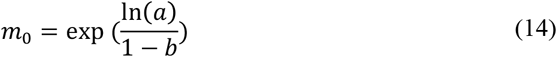

Ma (2015) extended TPL to the community level, introducing Taylor’s Power Law Extensions (TPLE). TPLE parameters can be used to calculate the community heterogeneity threshold (CHT), which serves as the community-level analog to the population-level PACD. This framework provides insights into spatial heterogeneity and aggregation patterns within microbial communities.

## Results

### 1. Sloan’s Near-Neutral Model (SNM) and Fisher’s Exact Test

We applied Sloan’s near-neutral model (SNM) to analyze the gut microbiome datasets associated with Parkinson’s disease (PD). The model was fitted by treating each sample site as both the source and destination community. The results, presented in **Table S2**, revealed high goodness-of-fit (R^2^ = 0.477 for healthy and 0.532 for diseased communities), indicating that the SNM effectively captured the ecological dynamics of the microbiota. In healthy communities, 40.0% of species were classified as neutral, while 47.4% and 12.6% were positively and negatively selected, respectively. **Fig 1** shows the species distribution of the gut microbiome associated with Parkinson’s disease fitting the SNM in healthy communities. In diseased communities (see Fig 2), the proportion of neutral species decreased to 38.1%, with 45.4% positively selected and 16.5% negatively selected species.

**Fig 1.**
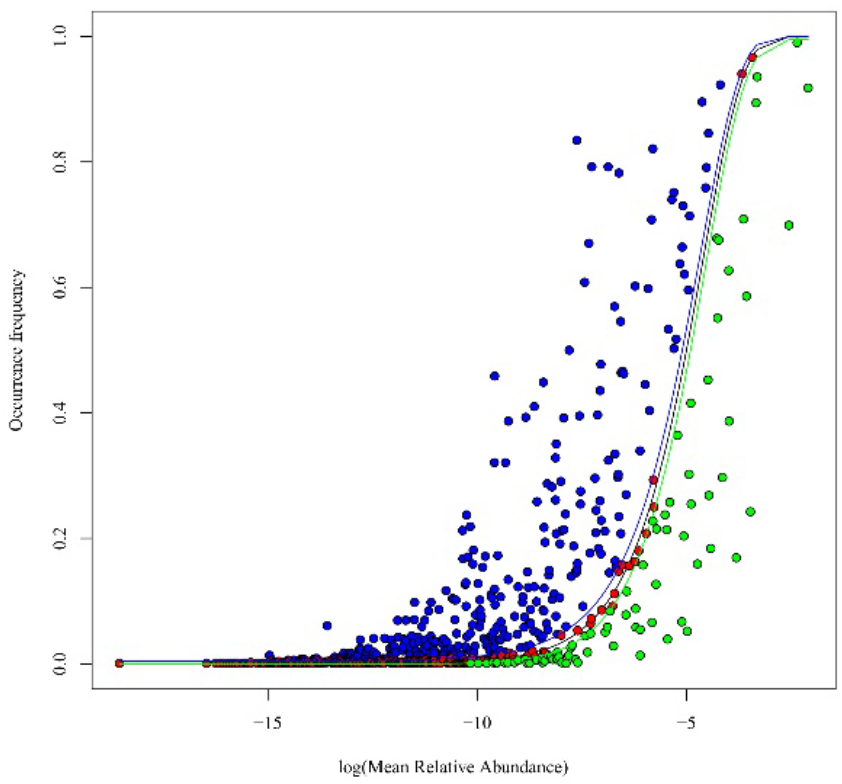
Fitting Sloan et al. (2006, 2007) near neutral model to the gut microbiomes associated with Parkinson’s disease. Both Source Community and Destination Community are the same healthy group. The blue nodes represent positively selected species, the red nodes represent neutral species and the green nodes represent negatively selected species.

**Fig 2.**
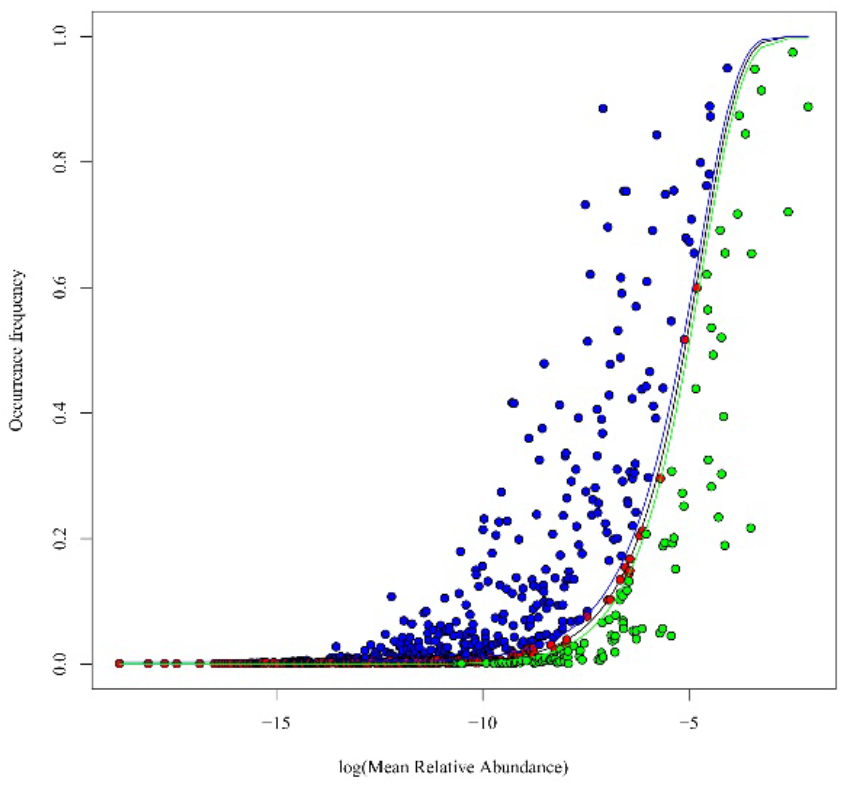
Fitting Sloan et al. (2006, 2007) near neutral model to the gut microbiomes associated with Parkinson’s disease. Both Source Community and Destination Community are the same healthy group. The blue nodes represent positively selected species, the red nodes represent neutral species and the green nodes represent negatively selected species.

Fisher’s exact test (**Table S3**) was used to compare the species categories between healthy (H) and diseased (D) treatments. No significant differences were detected, as indicated by odds ratios close to 1 (range: 0.953–1.310) and P-values > 0.05. This suggests that PD does not significantly alter the preference for neutral or selected species.

### 2. Normalized Stochasticity Ratio (NSR) Analysis

The normalized stochasticity ratio (NSR) was calculated to quantify the neutrality levels of species categories (**Table S4**). PD significantly increased the neutrality level, as evidenced by higher NSR values for neutral species (healthy: 0.875; diseased: 1.000; P < 0.001). Conversely, both positively and negatively selected species exhibited lower NSR values in PD compared to healthy conditions (positively selected: 0.316 vs. 0.299; negatively selected: 0.534 vs. 0.495; P < 0.001), indicating reduced neutrality (or higher selection levels) in diseased states. Across all species, the healthy treatment demonstrated higher neutrality and lower selection levels compared to the diseased treatment.

### 3. Shared Species Analysis (SSA)

Shared Species Analysis (SSA) revealed significant compositional shifts in all species categories due to PD (**Table S5**). The observed shared OTUs were significantly lower than expected under randomization (P < 0.05), indicating that PD altered the species composition across neutral, positively selected, and negatively selected categories. Further analysis identified species unique to PD (PD-H), including 149 neutral, 52 positively selected, and 57 negatively selected species (**Table S6**).

### 4. Dominance network Analysis

Microbial correlation networks were constructed using Spearman’s rank correlation (P < 0.05, FDR-corrected) to evaluate interactions within each species category (Table S7). In healthy communities, the positive-to-negative edge ratio (P/N ratio) was highest for neutral species (5,469), followed by positively selected (8.654) and negatively selected species (8.19). A similar trend was observed in PD, with neutral species exhibiting the highest P/N ratio (121.513). Network density was highest for positively selected species in both healthy (131.993) and diseased (142.857) conditions, indicating stronger interactions within this category.

Figures 3 and 4 illustrate the network graphs of the three species categories classified by SNM modeling analysis for healthy and PD treatments, respectively. A notable property is that neutral networks displayed significantly fewer negative links (antagonistic interactions), consistent with the species equivalence principle of neutral theory. This property is evident not only through visual inspection of Figures 3 and 4 but also through the P/N ratio, which is significantly larger for neutral networks, as previously discussed. These findings underscore the distinct interaction patterns of neutral species, characterized by reduced antagonism and enhanced cooperative interactions. In Figures 3 and 4, positive links were omitted to emphasize the distribution of negative links. Since the positive-to-negative edge ratios (P/N ratios) are provided, no information is lost by this omission, as the number of positive links can be easily inferred from the P/N ratios and the counts of negative links. This approach allows for a clearer visualization of antagonistic interactions while retaining the ability to reconstruct the full network structure.

**Fig 3.**
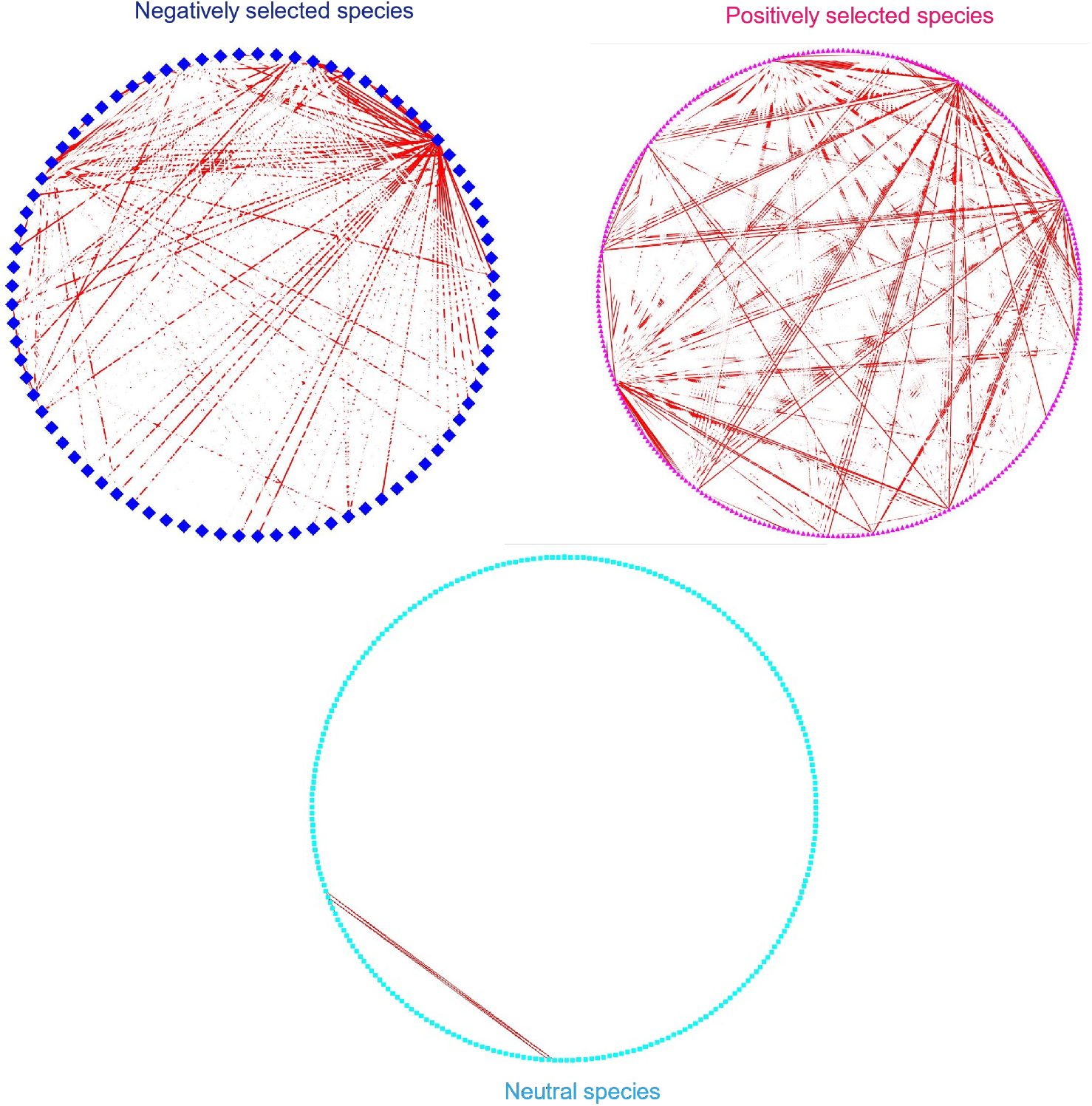
The network for three species group in SNM (Stochastic Niche Neutral Model) of the healthy group. For the Spearman’s rank correlation, the dominance values of OTUs were utilized to construct the microbial correlation network (*P*<0.05 and after FDR). The magenta triangle nodes represent positively selected species, the cyan round nodes represent neutral species and the dark blue diamond nodes represent negatively selected species. Edges in red represent negative correlations; edges in white represent positive correlations. For this figure we only show the negative correlations for species inside each species group.

**Fig 4.**
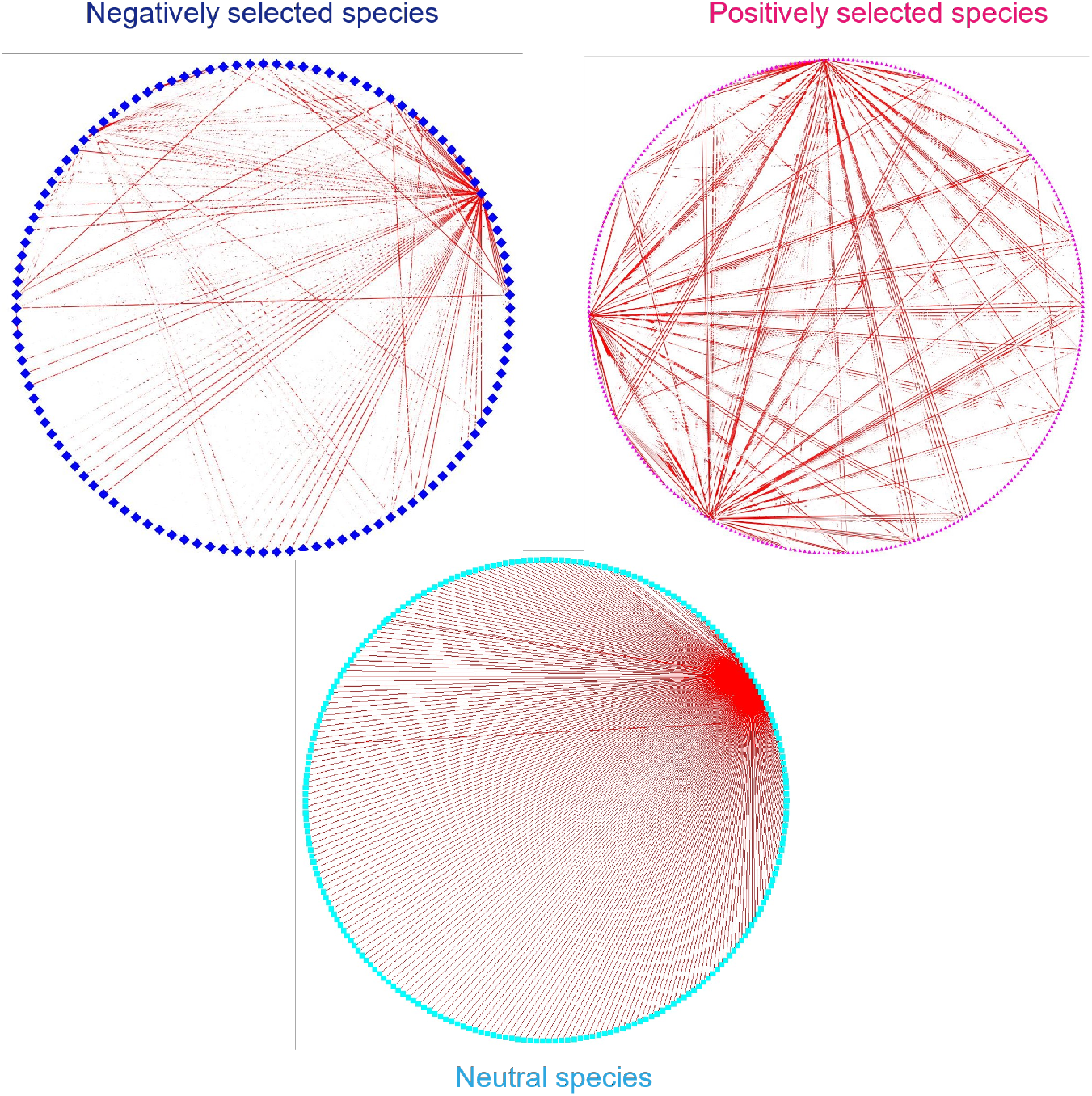
The network for three species group in SNM (Stochastic Niche Neutral Model) of the PD group. For the Spearman’s rank correlation, the dominance values of OTUs were utilized to construct the microbial correlation network (*P*<0.05 and after FDR). The red triangle nodes represent positively selected species, the cyan round nodes represent neutral species and the dark blue diamond nodes represent negatively selected species. Edges in red represent negative correlations; edges in white represent positive correlations. For this figure we only show the negative correlations for species inside each species group.

## 5. Relative Abundance and Dominance

The relative abundance of species categories exhibited the following trend: negatively selected > positively selected > neutral (Fig. 5). However, this pattern represents a general trend rather than a strict hierarchy, as the species ranks within these categories were intermixed rather than distinctly ordered.

**Fig 5.**
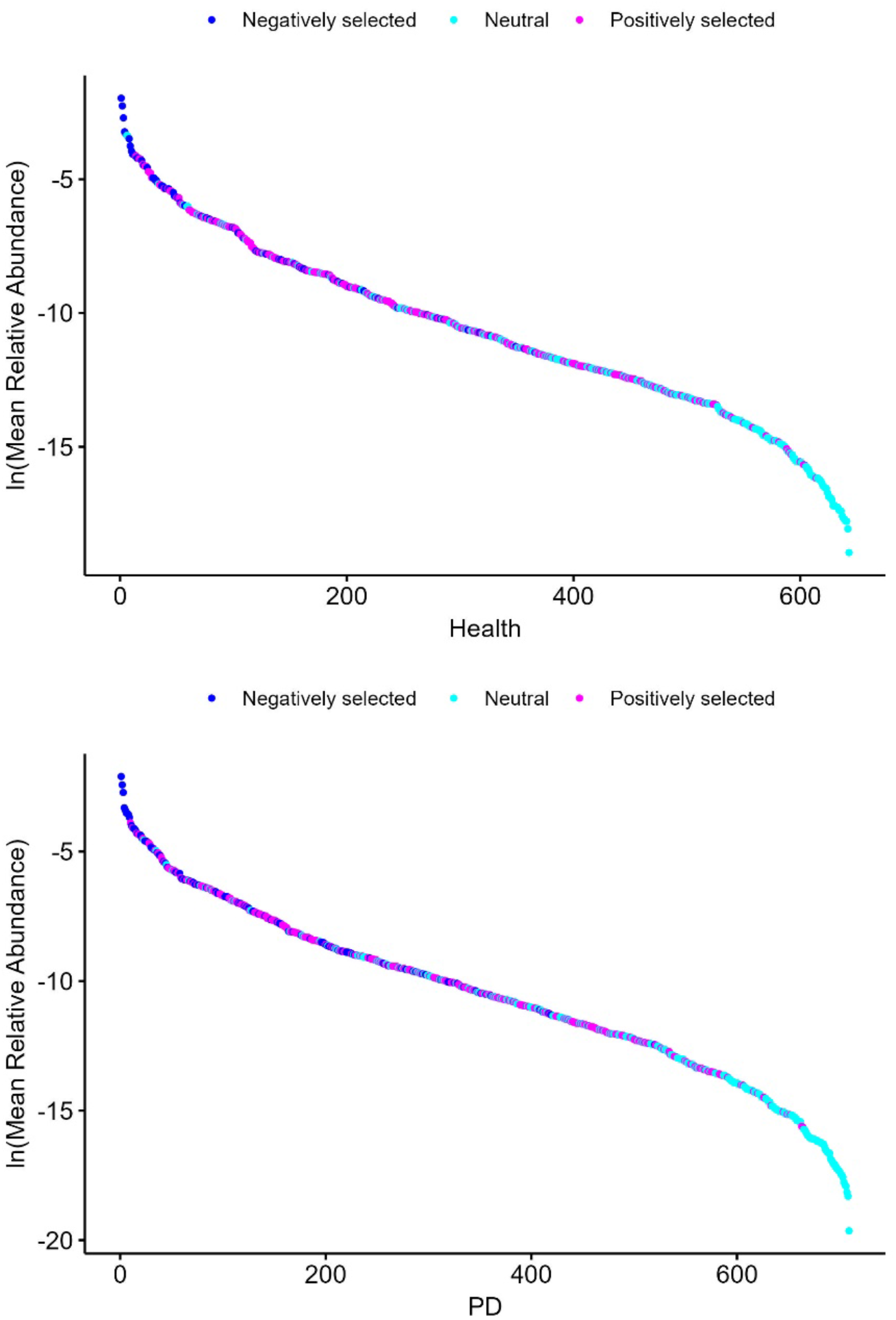
The relative abundance rank curve for microbial species to the gut microbiomes associated with Parkinson’s disease. The cyan nodes represent neutral species, the blue nodes represent negatively selected species, the magenta represent positively selected species.

This trend was consistent across both healthy and diseased groups, indicating that negatively selected species dominate the microbiota irrespective of disease status.

### 6. The Community Spatial Heterogeneity for each species category in SNM

Table S8 presents the fitting of Taylor’s Power Law Extensions (TPLE) to each species category classified by Sloan’s SNM model. In the healthy group, the *b*-values were 1.899 for neutral species and 1.864 for selected species, while in the PD group, these values were 1.886 and 1.888, respectively. The similarity in *b*-values across both health statuses indicates that the scaling of spatial heterogeneity remains consistent between neutral and selected species. This suggests that heterogeneity-scaling is invariant across healthy and diseased treatments.

## Conclusions and Discussion

The results presented above demonstrate a coherent narrative regarding the influence of Parkinson’s disease (PD) on the stochasticity of gut microbiomes. The first analysis, utilizing Sloan’s Near-Neutral Model (NNM) and Fisher’s exact tests, revealed that the NNM fitted the gut microbiome data satisfactorily. However, the proportions of the three species categories— neutral species, positively selected species, and negatively selected species—remained unchanged between PD and healthy conditions, suggesting no significant shift in the overall structure of species categories due to PD.

The second analysis, based on the Normalized Stochasticity Ratio (NSR), indicated that PD is associated with higher selection levels (or lower neutrality levels). This suggests a shift in stochasticity levels due to PD, which appears inconsistent with the findings from Sloan’s NNM and Fisher’s exact tests. We postulate that this inconsistency arises because Sloan’s NNM and Fisher’s tests only capture one type of holistic difference—the proportions of species categories—while the NSR reveals another type of holistic difference, reflecting changes in the underlying stochasticity and selection pressures.

The third analysis, using Shared Species Analysis (SSA), supports this postulation. SSA revealed that while the proportions of neutral, positively selected, and negatively selected species remained constant, the compositional makeup (i.e., the specific memberships) of these categories was significantly altered by PD (P < 0.05). This indicates that although the overall structure of species categories is preserved, the individual species within each category are reshuffled due to the disease.

The fourth analysis, involving network analysis, demonstrated that the network of neutral species contains fewer antagonistic interactions (negative links) compared to positively and negatively selected species, as evidenced by a higher positive-to-negative edge ratio (P/N ratio). This is consistent with the expectation that neutral species, being ecologically equivalent, exhibit more cooperative (positive) interactions and fewer competitive (negative) interactions.

The fifth analysis demonstrated that neutral species typically display lower relative abundances compared to positively and negatively selected species, highlighting their distinct ecological roles and dynamics within the gut microbiome. Additionally, TPLE modeling revealed that heterogeneity-scaling remains consistent across species categories and disease status, indicating an invariant pattern of spatial heterogeneity.

## Data Availability

All data reanalyzed in this study are available in public domain.

